# SARS-CoV-2 lineage-specific disease symptoms and disease severity in São Caetano do Sul city, Brazil

**DOI:** 10.1101/2023.10.19.23297252

**Authors:** Flavia Cristina da Silva Sales, Carlos Augusto Prete, Leandro Abade, Lewis Fletcher Buss, Darlan da Silva Candido, Ingra Morales Claro, Filipe Romero Rebello Moreira, Erika Regina Manulli, Ligia Capuani, Camila Alves da Silva Maia, Beatriz Araujo Oliveira, Thais Coletti, Heuder Gustavo Oliveira Paiao, Silvia Figueiredo Costa, Maria Cassia Mendes Correa, Fabio Eudes Leal, Kris Varun Parag, Vítor Heloiz Nascimento, Nuno Rodrigues Faria, Ester Cerdeira Sabino

## Abstract

**Background:** The city of São Caetano do Sul, Brazil, established a web-based platform to provide primary care to suspected COVID-19 patients, integrating clinical and demographic data and sample metadata. Here we describe lineage-specific spatiotemporal dynamics of infections, clinical symptoms, and disease severity during the first year of the epidemic.

**Methods:** We selected and sequenced 879 PCR+ swab samples (8% of all reported cases), obtaining a spatially and temporally representative set of sequences. Daily lineage-specific prevalence was estimating using a moving-window approach, allowing inference of cumulative cases and symptom probability stratified by lineage using integrated data from the platform.

**Results:** Most infections were caused by B.1.1.28 (41.3%), followed by Gamma (31.7%), Zeta (9.6%) and B1.1.33 (9.0%). Gamma and Zeta were associated with larger prevalence of dyspnoea (respectively 81.3% and 78.5%) and persistent fever (84.7% and 61.1%) compared to B.1.1.28 and B.1.1.33. Ageusia, anosmia, and coryza were respectively 18.9%, 20.3% and 17.8% less commonly caused by Gamma, while altered mental status was 108.9% more common in Zeta. Case incidence was spatially heterogeneous and larger in poorer and younger districts.

**Discussion:** Our study demonstrates that Gamma was associated with more severe disease, emphasising the role of its increased disease severity in the heightened mortality levels in Brazil.

## Introduction

Understanding the natural history of a new disease is crucial during an epidemic, but establishing large cohorts can be challenging in emergency situations. During the COVID-19 pandemic some studies leveraged existing infrastructure to understand the clinical manifestation of the disease in their community (1), while others described hospital cohorts (2–4).

In April 2020, a digital platform was created in the Municipality of São Caetano do Sul (SCS) in the metropolitan region of Sao Paulo city to assist all suspected coronavirus disease 2019 (COVID-19) patients and collect clinical data using standardised questionnaires. This initiative was made possible through the collaboration of the local Health Department, the University of São Caetano do Sul, and the University of São Paulo. This platform enabled systematic testing of all suspected cases and monitoring of confirmed severe acute respiratory syndrome coronavirus 2 (SARS-CoV-2) cases in the community (5)..

This allowed us to provide a detailed description of the evolution of the epidemic in the city during the first year of the epidemic in Brazil, which covers the emergence and spread of the yet poorly understood Gamma variant of concern (VOC), and the impact of this VOC on the affected population’s clinical manifestation. The Gamma VOC was first detected in Manaus, a region that has achieved a high attack rate during the first wave (6,7) and quickly expanded to all Brazilian cities causing a dramatic increase of COVID-19 deaths. Although the SARS-CoV-2 Gamma VOC wave resulted in a more deadly epidemic when compared to the first wave in Brazil, little is known about the differences in clinical manifestations and disease severity of this lineage in relation to previously circulating strains, including its ancestral lineage B.1.1.28 (6,8).

In this paper, we generate a dataset containing integrated epidemiological, clinical and genomic data and characterise the spatiotemporal dynamics, clinical symptoms and disease severity of the main lineages circulating in São Caetano do Sul during the first year of the SARS-CoV-2 epidemic.

### Study area description

São Caetano do Sul (SCS) is a municipality located in the metropolitan region of São Paulo, Brazil, with a population of 162.763 inhabitants (9). It is intensely conurbated with the city of São Paulo, Santo André and São Bernardo do Campo. The municipality has a high Human Development Index (HDI) of 0.862, with a low illiteracy rate of 1.5% compared to an average of 4.2% in the state of São Paulo.

### Corona Sao Caetano primary care programme

The Corona São Caetano program (https://coronasaocaetano.org) was an initiative implemented on April 6, 2020 designed to organise the public health response to the COVID-19 pandemic. The program instructed residents to register through an online platform or telephone contact to receive a telehealth consultation for a risk assessment. Pregnant women or patients showing alarming symptoms such as shortness of breath, persistent fever, confusion, or lethargy were advised to seek medical attention at the hospital. Patients without risk were instructed to stay at home and perform self-collection of samples for diagnosis, receiving telehealth services for up to 14 days after the onset of symptoms. (5).

## Materials and Methods

### Ethical approval

This study was approved by the National Research Ethics Committee under protocol number CAAE 30127020.0.0000.0068 and 32424720.8.0000.0068. The committees waived the need for informed consent and allowed the development of an unidentified analytical dataset for analysis.

### Representative selection of samples for sequencing

The São Caetano platform routinely collects patient addresses and identifies the corresponding neighbourhood among 15 neighbourhoods in the city. To obtain a comprehensive spatial representation of positive cases across the city, we randomly selected two positive samples per epidemiological week and per neighbourhood. This approach allowed us to include 1063 positive cases, covering the period from April 6, 2020 to April 30, 2021, and ensured that we captured the diversity of cases from different locations in the city.

### SARS-CoV-2 diagnosis

All samples were tested by PCR using ALTONA RealStar® SARS-CoV-2 RT-PCR 1.0 Kit (Hamburg, Germany), Mico BioMed RT-qPCR Kit (Seongnam, South Korea) according to the manufacturer’s instructions and kit availability.

### Whole genome sequencing

To sequence the SARS-CoV-2 positive samples, we used a tilling-amplicon multiplex PCR technique using V3 or V4 scheme as previously described (10); (8) (11); (12). Sequencing libraries were generated using the SQK-LSK109 Kit, were loaded onto an R9.4.1 flow-cell on the MinION device and sequenced using MinKNOW 22.3.6 (ONT, UK).

### Bioinformatic analysis

The FAST5 files generated during sequencing were basecalled, demultiplexed, and trimmed using Guppy software version 6.0.7 (Oxford Nanopore Technologies, UK). To obtain consensus sequences, FASTQ files were mapped against the reference genome of SARS-CoV-2 isolate Wuhan-Hu-1 (GenBank accession number MN908947) using the Minimap2 program version 2.28.0 (13) and SAMTools (14) converting the files into BAM format. Length filtering and quality testing were performed for each barcode using ARTIC guppyplex (https://artic.network/ncov-2019/ncov2019-bioinformatics-sop.html). Genome regions with a depth of <20-fold were not included in final consensus sequences, and these positions are represented with N characters. Sequences of low quality, less than 75 percent genome coverage or that presented contamination were discarded from final analysis. Lineages were classified using the Pangolin COVID-19 Lineage Assigner software tool (http://pangolin.cog-uk.io/) and Nextclade (https://clades.nextstrain.org/). Consensus sequences were submitted to the GISAID platform.

### Epidemiological and clinical data

To conduct epidemiological and clinical analyses, we extracted data from the Corona São Caetano platform for confirmed SARS-CoV-2 cases. Non-identifiable clinical data including demographic information (age, gender, education level and neighbourhood) and clinical information such as symptom onset date and reported symptoms were collected through a standardised questionnaire.

The SCS epidemic can be divided into three phases that comprise: Phase 1: between April 4 and September 30, 2020, during which no variants of concern (VOCs) or variants of interest (VOIs) were detected; Phase 2 comprising the period between October 1, 2020 and December 31, 2020, marked by the emergence of Zeta VOI (P.2); and finally, Phase 3 spanning from January to April 2021, characterised by the widespread circulation of the Gamma VOC (P.1) in Brazil.

### Inference of lineage prevalence over time

We estimated the daily prevalence of each lineage by applying a moving window of variable size to the daily number of sequenced samples identified as that lineage. At each instant, only samples contained in the moving window are considered in the calculation of the prevalence. Our algorithm uses a larger window size in periods where few samples are sequenced, avoiding excessive noise, and decreases the window size in periods with large number of sequences, increasing time precision. We chose to use a time-varying window because the crude prevalence of lineages B.1.1.28, B.1.1.33 and Zeta had multiple prevalence surges (see **Fig. 1S**), hence multinomial logistic regressions as used in (15) cannot be employed to infer the continuous-time prevalence.

Let *S*[*n*, ℓ] be the number of sequenced samples associated with lineage ℓ ∈ *A* at day *n*, where *A* = {Gamma, Zeta, B. 1.1.28, B. 1.1.33, Other}. All lineages that are not classified as Gamma, Zeta, B.1.1.28 or B.1.1.33 are assigned into a single group ‘Other’. Note that *S*[*n*, ℓ] is a sparse signal for a fixed ℓ, being different than zero only in days where a sequenced PCR+ sample is identified as belonging to lineage ℓ.

Define for each day *n* ≥ 1 the window radius *L*[*n*] as the smallest *L*′ ≥ *L*_*min*_ such that

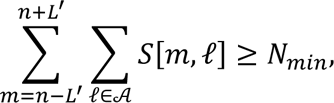

where *L*_*min*_ and *N*_*min*_are predefined parameters representing respectively the smallest window radius and number of samples allowed. These quantities are arbitrary, and in this work we chose *L*_*min*_ = 7 days and choose *N*_*min*_ as the ceiling of the average number of samples contained in a window of radius *L*_*min*_ = 7 days (and length 2*L*_*min*_ + 1 = 15 days). Hence, 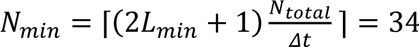, where *N* = 879 is the total number of sequences in the period and *Δt* = 391 days is the total duration of the study. Hence, this method defines a window whose time-varying radius cannot be smaller than 7 days and increases until a minimum of 34 samples are included in the window.

Once *L*[*n*] is computed, the prevalence of each lineage ℓ at instant *n*, denoted by *ρ*[*n*, ℓ], is obtained by calculating the crude prevalence of the samples in the interval [*n* − *L*[*n*], *n* + *L*[*n*]]:

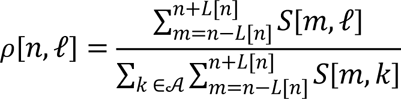

### Effective reproduction number estimation

We used the daily number of cases reported in the CSC Platform and the estimated daily prevalence *ρ*[*n*, ℓ] to infer lineage-specific effective reproduction number (*R*_*t*_) estimates. For that, we used the algorithm EpiFilter presented in (16) with step *η* = 0.1. We used a log-normal distribution with log-mean 1.09 and log-standard deviation 0.72 as generation interval distribution based on Brazilian serial intervals measured in the early phase of the epidemic in Brazil (17).

To estimate lineage-specific *R*_*t*_ and its confidence intervals, we calculated the daily lineage-specific incidence *I*[*n*, ℓ] as *I*[*n*, ℓ] = round(*ρ*[*n*, ℓ]*I*[*n*]), where *I*[*n*] = ∑_ℓ∈*A*_ *I*[*n*, ℓ] is the measured total incidence at day *n* (that is, the number of PCR+ individuals that had their first symptoms at day *n*) and round(*x*) is the nearest integer to *x*. For each ℓ ∈ *A*, we apply EpiFilter (16) using *I*[*n*, ℓ] as input, obtaining an estimate of *R*_*t*_ for lineage ℓ. We also estimated *R*_*t*_ with no disaggregation by lineage by simply running EpiFilter using the daily number of cases.

In addition to Rt, the estimates of *I*[*n*, ℓ] also allowed us to infer the cumulative number of cases *I*_*C*_[*n*, ℓ] caused by each lineage as

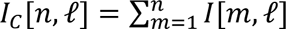

### Estimation of Symptom Probabilities

We considered that a patient with a sequenced PCR+ result infected by a given lineage had a given symptom if the symptom was reported in any of the medical visits. Credible intervals were calculated using a Bayesian approach assuming a uniform prior distribution for the probability of having the given symptom *p*_*S*_ in the interval [0,1]. The likelihood is *N*^+^|*p*_*s*_ ∼ Binomial(*N*, *p*_*s*_), where *N* is the total number of sequenced PCR+ patients infected by that lineage and *N*^+^is the subset of this group that had the given symptom. Therefore, the posterior distribution is *p*_*s*_|*N*^+^ ∼ Beta(1 + *N*^+^, 1 + *N* − *N*^+^). The quantiles of this Beta distribution are drawn to estimate median and 95% credible intervals.

To validate our results, symptom probabilities were also estimated considering all PCR+ patients, not only patients with sequenced samples. For that, we imputed the lineage that infected each patient based on the inferred lineage prevalence *ρ*[*n*, ℓ] on the day of symptom onset. The procedure used to obtain the posterior distribution for *p*_*s*_ is the same as above, except that *N*^+^ and *N* are estimated by 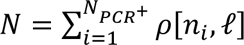 and 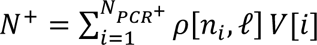, where *N*_*PCR*+_ is the number of PCR+ patients, *n*_*i*_ is the date of symptom onset for patient *i* and *V*[*i*] = 1 if patient *i* had the given symptom, else *V*[*i*] = 0.

### P-values and probability of more frequent symptoms

We performed hypothesis tests to determine whether a given symptom is more or less frequently caused by Gamma or Zeta when compared to lineages B.1.1.28 and B.1.1.33. For that, we merged infections caused by B.1.1.28 and B.1.1.33 into a single class “B.1.1.28 or B.1.1.33” and we used the exact Fisher test with confidence level of 95% to determine if the prevalence of that symptom is significantly different for Gamma or Zeta when compared to B.1.1.28 and B.1.1.33. Two hypothesis tests were performed for each symptom (one for Gamma and one for Zeta), resulting in 34 tests. Due to the large number of tests, we performed a Benjamini-Hochberg correction with a false positive rate of 10% which rejected all hypothesis tests with p ≤ 0.0033.

To better interpret the p-values obtained with the hypothesis tests, we also estimated the probability of a given symptom being more frequently caused by Gamma or Zeta when compared to B.1.1.28 and B.1.1.33. For that, we generated *N*_*samples*_ = 10,000 samples from the posterior distribution of each symptom prevalence and calculated the empirical probability of the symptom being more frequent in Gamma or Zeta when compared to B.1.1.28 and B.1.1.33. Therefore, denoting as *p*_1_, *p*_2_, …, *p*_*Ns*_ and *r*_1_, *r*_2_, …, *r*_*Ns*_ respectively posterior samples of Gamma or Zeta and B.1.1.28 or B.1.1.33, the probability of that symptom being more frequent in patients infected by Gamma or Zeta is 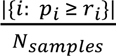, where |{*i*: *p*_*i*_ ≥ *r*_*i*_}| is the number of indices *i* such that *p*_*i*_ ≥ *r*_*i*_.

### Estimation of Case Fatality and Hospitalisation Rates

Case Fatality Rate (CFR), Case Hospitalisation Rate (CHR) and In-hospital Fatality Rate (HFR) were estimated using cases reported in CSC platform until March 31, 2021, and Severe Acute Respiratory Infection (SARI) hospitalisations reported in SIVEP-Gripe. The latter is an open dataset that contains individual-level information such as city of hospitalisation, city of residence, date of symptom onset and outcome (that is, recovery or death). This dataset is available at https://opendatasus.saude.gov.br/.

We selected SARI patients that lived in São Caetano do Sul and were hospitalised in the state of São Paulo. We calculated the daily number of cases and deaths using the date of symptom onset, available in both datasets.

As in the procedure used to estimate symptom probabilities, we used a Bayesian approach to infer fatality and hospitalisation rates assuming a uniform distribution in the interval [0,1] for the probability of severe or fatal outcome *p*_*S*_. Therefore, the posterior distribution for the fatality or hospitalisation rate in a given period is

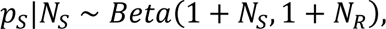

where *N*_*S*_ is the number of observed severe or fatal outcomes and *N*_*R*_ is the observed number of recoveries. Therefore, *N*_*S*_ is the number of hospitalisations for CHR calculation, and the number of deaths for HFR and CFR estimates, while *N*_*R*_ is the number of SARI recoveries for HFR inference and the difference between the number of PCR+ cases and the number of deaths for CFR and CHR calculations.

## Results

### Epidemiological context

From April 6, 2020 to April 30, 2021, the Municipal Bulletin of SCS notified 10,880 cases of COVID-19. During the same period, the Corona São Caetano platform included 38,733 suspect cases on its digital platforms. From these, 26,584 tests for SARS-CoV-2 were applied, resulting in 6,905 positive cases that represents 63% of all notified cases in the city. Of these positive cases, 1,063 samples (15.4%) were selected for sequencing, and 879 sequences were obtained with more than 75x coverage (**Fig. 1**).

**Figure 1.**
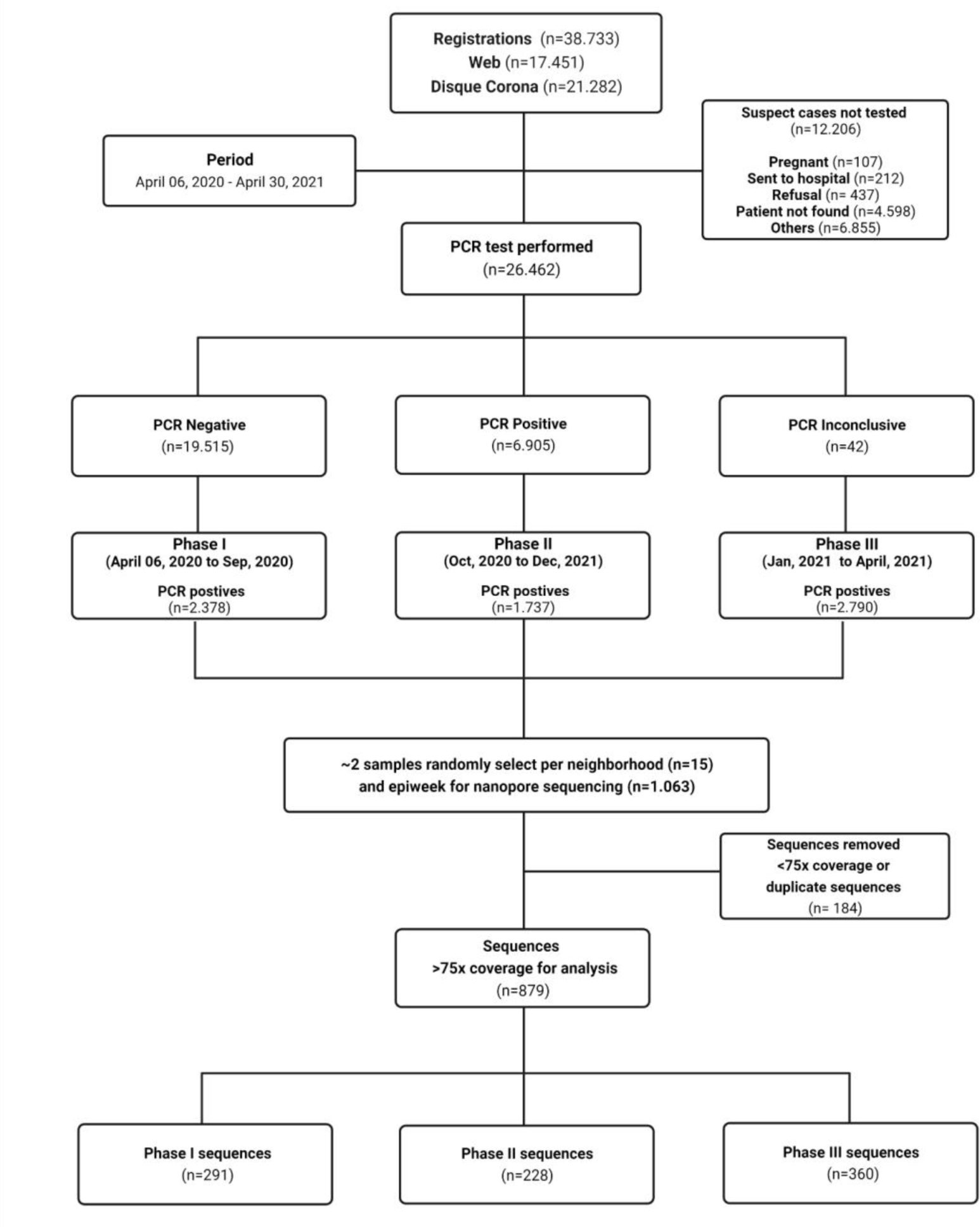
Flowchart containing information about the study design. 38,733 patients registered on the CSC Platform for care. Of the 26,462 individuals who self-collected for the diagnosis of SARS-CoV-2, a total of 6,905 individuals were positive, 19,515 were negative, and 42 were inconclusive. From these positive samples, we randomly selected ∼2 positive samples per neighbourhood per epidemiological week for genome sequencing using the ARTIC protocol and the Oxford Nanopore sequencing platform. For epidemiological analyses and lineage assignment, we considered genomes with >75x coverage (n=879).

Because of our sampling strategy for sequencing, we obtained an epidemiologically, spatially, and temporally representative set of sequences. In **Table 1**, we compare positive and sequenced cases, and show that both groups had similar proportions of infection regardless of gender, age, and education. We performed a linear correlation analysis to compare PCR+ cases by neighbourhood and epidemiological week, considering sequences with >75x coverage (**Fig. 2S**). We obtained good correlation results both spatially (Pearson’s correlation coefficient = 0.87) and temporally (Pearson’s correlation coefficient = 0.45)

**Table 1.** Demographic and clinical characteristics of individuals from the Corona São Caetano program.

| Demographic and clinical characteristics of patients from the Corona São Caetano program |  |  |  |  |  |
| --- | --- | --- | --- | --- | --- |
| Characteristic | Total of positive cases (n=6905) |  | SARS-CoV-2 genome sequences (n=879) |  | p-value ** |
|  | Values | Freq (%) | Values | Freq (%) |  |
| **Sex** |  |  |  |  | 0.13 |
| Male | 2922 | 42.31% | 393 | 44.71% |  |
| Female | 3983 | 57.68% | 486 | 55.29% |  |
| **Age groups** | 12 ≤ 44 ≤ 105 |  | 12 ≤ 43 ≤ 92 |  | 0.4437 |
| 12–19 | 363 | 5.29% | 41 | 4.66% |  |
| 20–39 | 2618 | 37.94% | 329 | 37.43% |  |
| 40–59 | 2590 | 37.54% | 349 | 39.70% |  |
| 60+ | 1334 | 19.22% | 160 | 18.20% |  |
| **Educational level** |  |  |  |  | 0.6166 |
| Never attended school | 63 | 0.91% | 10 | 1.14% |  |
| Up to primary education | 1284 | 18.60% | 153 | 17.41% |  |
| High school | 3065 | 44.39% | 401 | 45.62% |  |
| University | 2467 | 35.73% | 308 | 35.04% |  |
| NA | 0 | 0.00% | 6 | 100.00% |  |
| **Essential occupation** |  |  |  |  | 0.7329 |
| Non-HCW essential job* | 1551 | 22.46% | 204 | 23.21% |  |
| Carers | 98 | 1.42% | 13 | 1.48% |  |
| HCW | 325 | 4.71% | 46 | 5.23% |  |
| No | 4904 | 71.02% | 609 | 69.28% |  |
| NA | 27 | 0.39% | 7 | 0.80 % |  |

NA= missing data;
\* Security, emergency services, supermarket, public transport and pharmacy workers
\*\* Pearson's Chi-squared test

In **Fig. 2**, we describe the evolution of the variants over time, the proportion of sequenced cases among confirmed COVID-19 cases and the total number of confirmed cases according to the CSC Platform and the Municipal Bulletin. B.1.1.28 and B1.1.33 co-circulated in the first wave, with cases decreasing by August 2020, but slowly rebounding when Zeta VOI started to spread. The first Gamma-infected case was detected in early January, and by the middle of February, this VOC became the predominant variant. Of the 879 sequences obtained, B.1.1.28, B1.1.33, Zeta and Gamma were the most common lineages detected in 26.2%, 8.7%, 11.7%, and 44.1%, respectively (**Table 1S**). Other variants were detected in the remaining 9.3% of cases. The most common lineages classified as ‘Other’ are: N.9 (23.4%), B.1.1 (18.2%), P.7 (13.0%) and the Alpha VOC (B.1.1.7) (9.1%).

**Figure 2.**
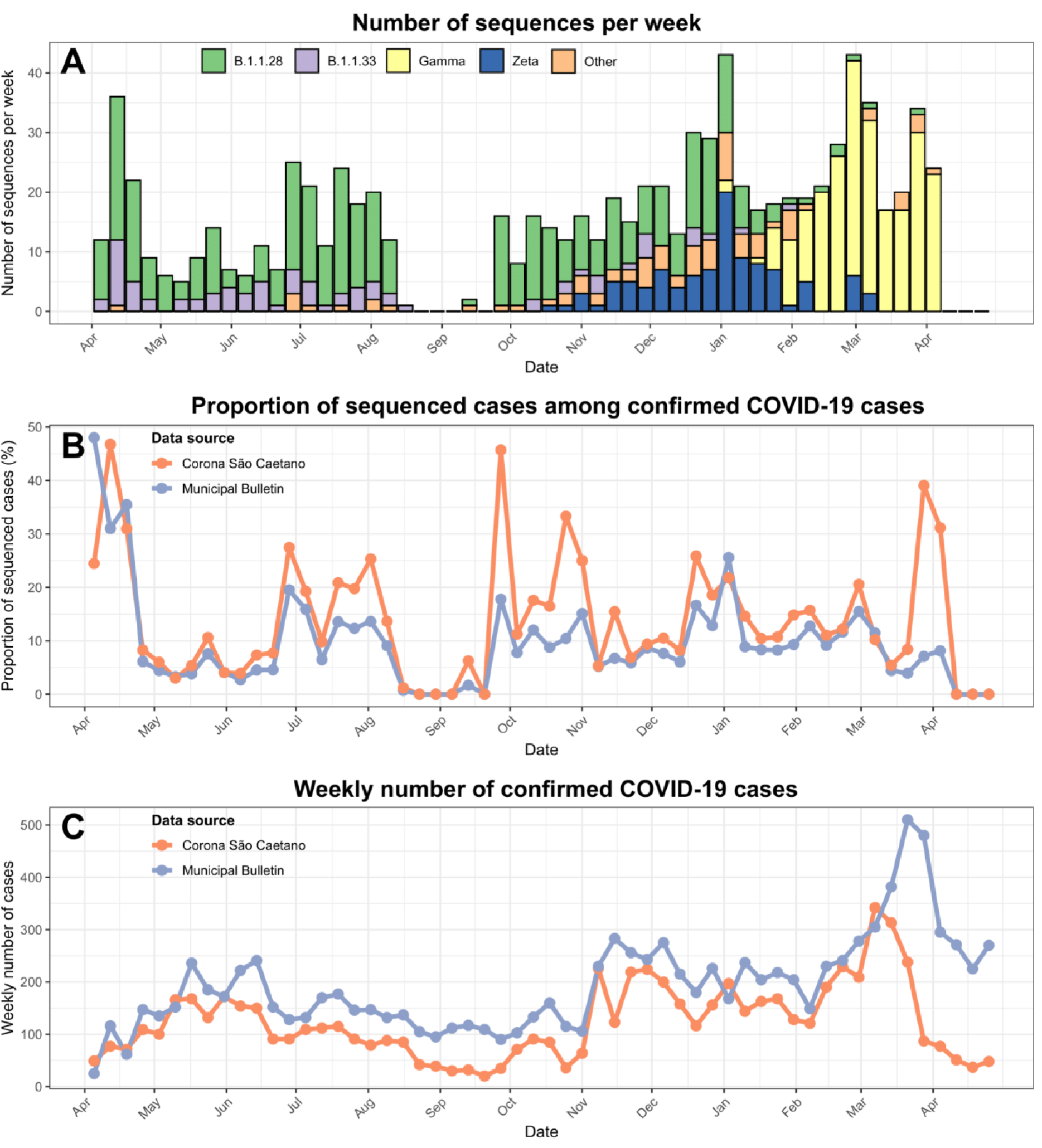
(A) Number of sequenced PCR+ samples by epidemiological week identified as the main ancestral lineages (B.1.1.28 and B.1.1.33), VOI (Zeta) and VOC (Gamma). Samples not identified as one of these lineages were assigned into the group ‘Other’. B) Proportion of sequenced PCR+ cases among all cases reported by the municipal bulletin (in blue) or Plataforma SC (orange). (C) Weekly number of confirmed cases in SCS according to Corona São Caetano (by date of symptom onset) and the municipal bulletin (by date of case notification).

We investigated the monthly and total incidence of COVID-19 cases by neighbourhoods within the city of SCS (**Fig. 3A** and **3B**). Incidence varied greatly across neighbourhoods, ranging from 34 to 69 cases per 1,000 inhabitants, and this spatial heterogeneity was consistent across time. **Fig. 3C** and **3D** shows the total incidence in districts in terms of average income per capita and proportion of population above 50 years old, demonstrating that neighbourhoods with lower per capita income and younger population had higher COVID-19 case incidence.

**Figure 3.**
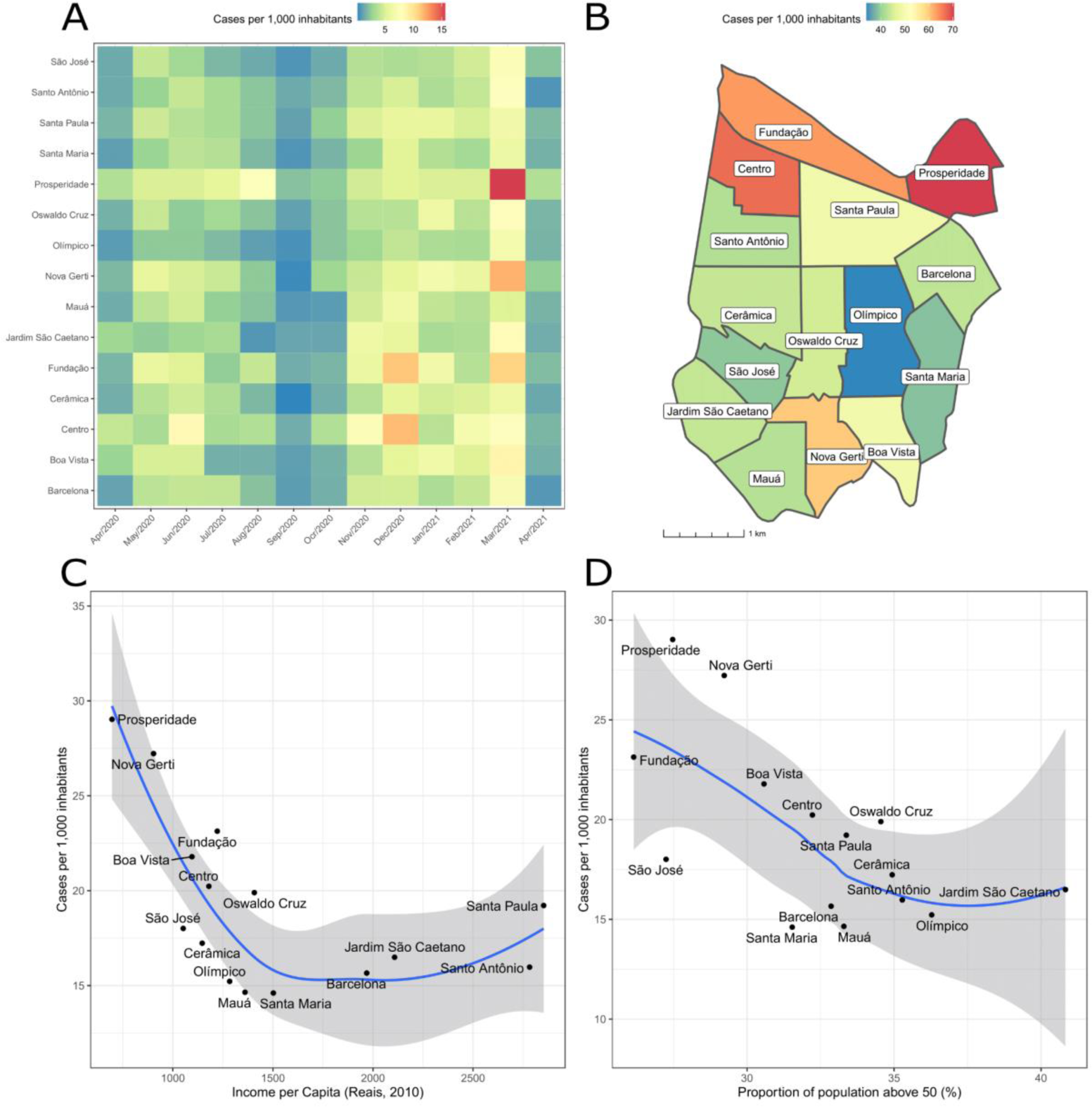
**A)** Heatmap showing the monthly incidence of SARS-CoV-2 by neighbourhood in São Caetano do Sul city between April 2020 - April 2021. Neighbourhoods are shaded according to the number of cases sampled. **B)** Map of total incidence of cases (n=6905) per 1000 inhabitants. Neighbourhoods are shaded according to the number of confirmed cases. **C and D)** Total incidence in each neighbourhood compared to the income per capita (C) and proportion of population above 50 years old (D). The curve in blue and ribbons in grey respectively represent the mean and 95% confidence intervals of a Loess regression with span=1.0.

The estimated daily and cumulative incidence for each lineage is shown in **Fig. 3S**. We infer that by April 30 2021, 41.3% of the cases were caused by B.1.1.28, followed by Gamma (31.7%), Zeta (9.6%) and B.1.1.33 (9.0%). Note that calculating the overall genomic prevalence using the estimated daily lineage-specific incidence leads to a more precise measurement than simply computing the proportion of sequenced cases, as the latter underestimates prevalence of lineages circulating during periods of high incidence.

Inferred lineage-specific incidences were used to calculate the effective reproduction number for the main strains that circulated during the studied period, shown in **Fig. 4**. During the first and second phases of the study, Rt fluctuated between 1 and 1.5 for both major strains (B.1.1.28 and B.1.1.33). In November 2020, a high Rt (>1.5) was observed for the B.1.1.28 lineage, coinciding with the emergence of the Zeta variant (formerly known as P.2). The Gamma VOC emerged during the circulation of the B.1.1.28 and Zeta lineage (6). In the first two months of the Gamma epidemic, an oscillation of the Rt between 1.0 and 1.5 was observed (**Fig. 4**). When comparing Rt during the first weeks after the first detected case of each lineage, Gamma’s Rt was similar to other lineages despite its increased transmissibility and reinfection capability (6), likely due to previous immunity.

**Fig. 4.**
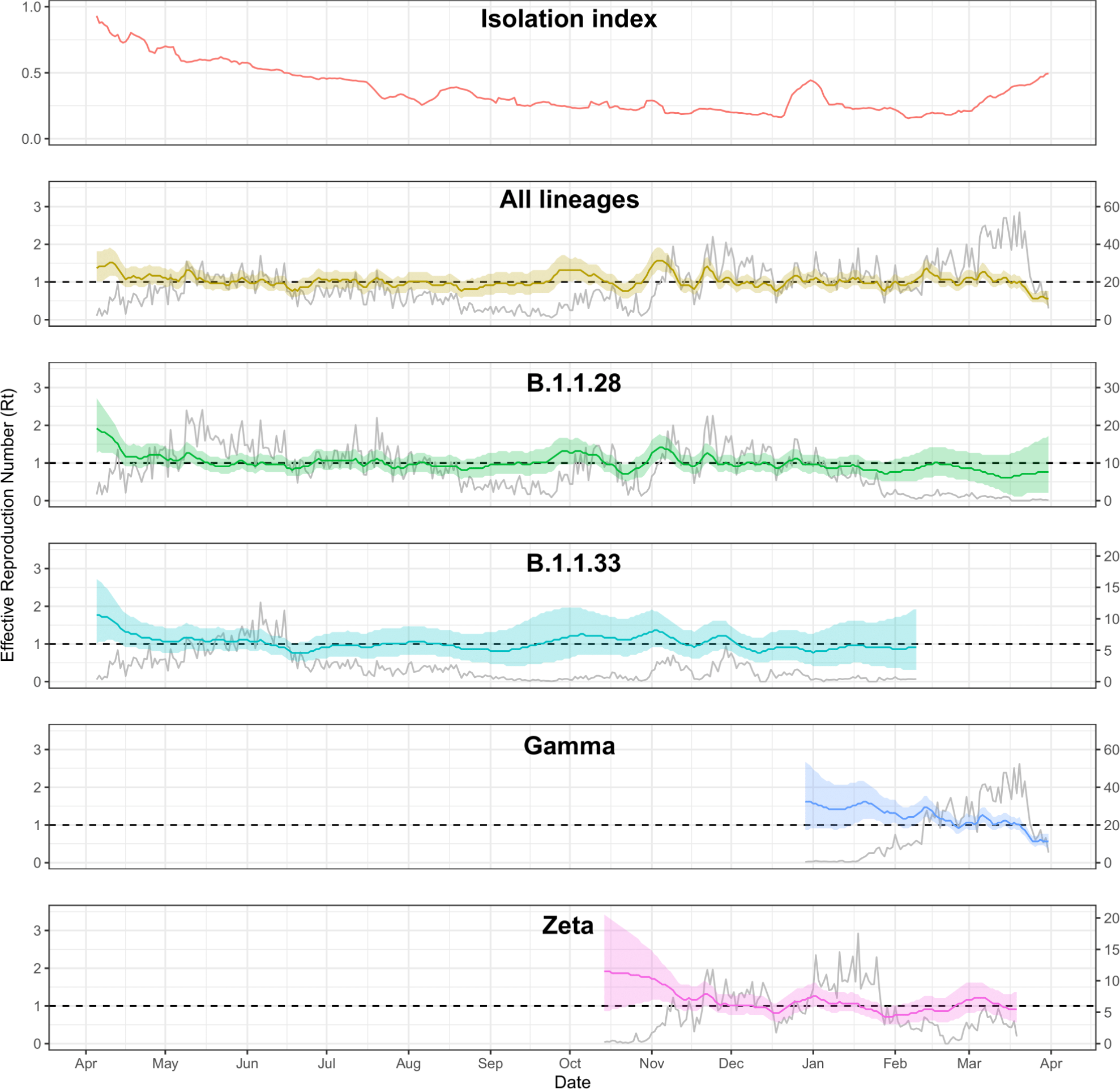
Lines show the median effective reproduction number (Rt) (y-axis on the left) estimated for each lineage, and the isolation index. Ribbons indicate 95% credible intervals. Grey curves represent the expected value of the estimated daily number of cases (y-axis on the right) by date of symptom onset caused by main lineages circulating between April/2020 and April 2021. Isolation index was extracted from the official website of the government of the state of São Paulo: https://www.saopaulo.sp.gov.br/coronavirus/isolamento.

During the whole period of study, overall and lineage-specific Rts oscillated around 1.0 with no large periods consistently below 1, with B.1.1.28 and B.1.1.33 having multimodal incidence patterns. This was likely due to continuous importation of cases from the neighbouring conurbated cities of São Paulo, Santo André and São Bernardo do Campo, in addition to a constantly decreasing isolation index until the Gamma-dominated wave in January 2021.

### Increased disease severity caused by Gamma VOC

Descriptive analyses of case fatality rate (CFR), case hospitalisation rate (CHR) and in-hospital fatality rate (HFR) were performed by crossing data from SIVEP-GRIPE and data from the Corona São Caetano platform. All three disease severity indicators were higher during the second epidemic wave (phase III) compared with the previous phases. (**Fig. 5**).

**Figure 5.**
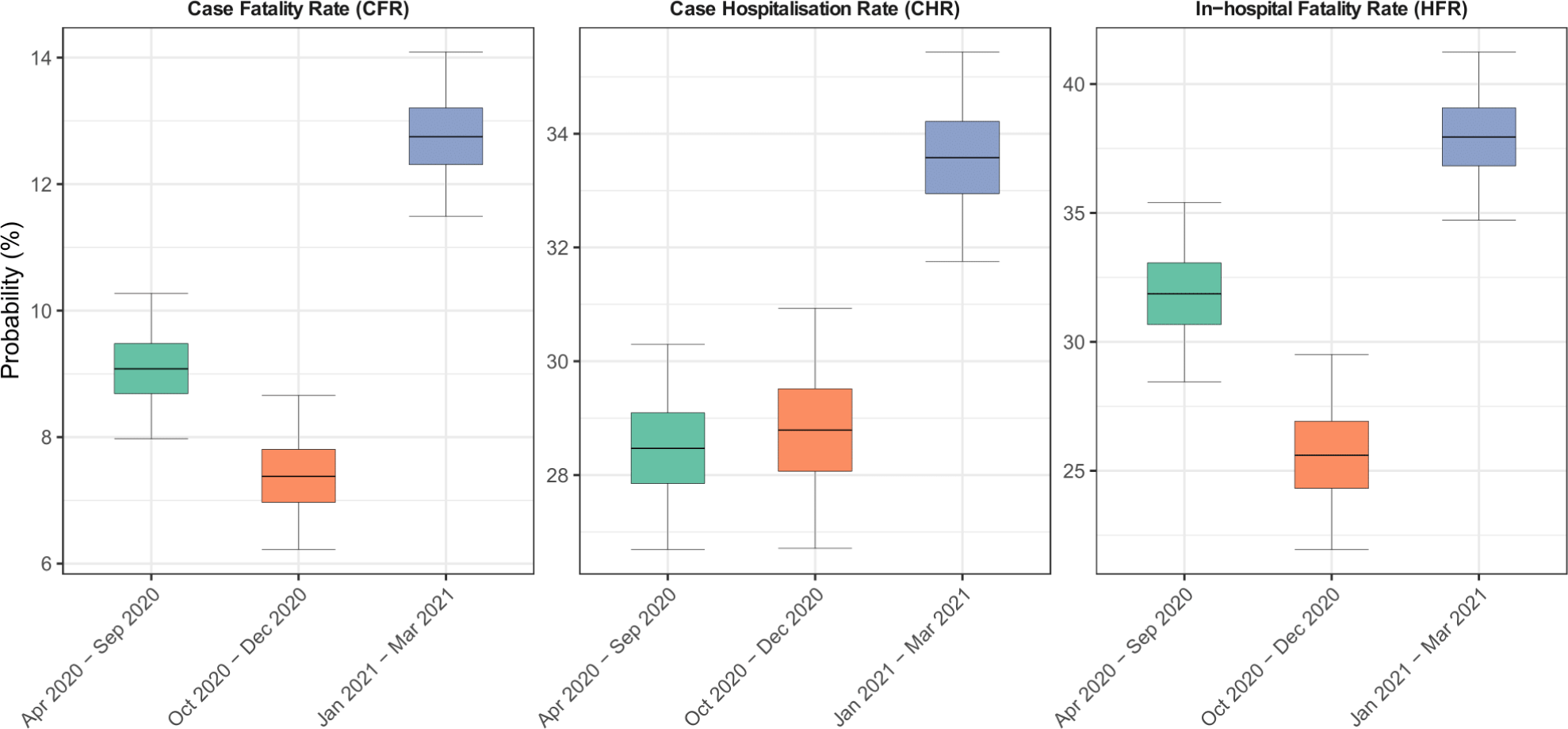
Inferred case fatality rate (CFR), hospitalisation rate (CHR) and in-hospital fatality rate (HFR) disaggregated by study period. All these indicators were higher during the third period when Gamma VOC was prevalent.

We performed multiple comparisons and plotted the proportion of each specific symptom according to the four most common lineages. We evaluated the 879 sequenced cases (**Fig. 6A**), and all 6,095 cases (**Fig. 6B**) after inputting the most probable variant according to the week of symptom initiation. Both plots show that symptoms related to more severe disease such as dyspnea and persistent fever were approximately two times more common among Gamma and Zeta lineages as compared to the ancestral variants (B1.1.28 and B1.1.33) (see **Table 2S** for exact values). Ageusia and anosmia was less common for the Gamma lineage and neurological symptoms were more common for the Zeta VOI. Due to the lack of limitation of sample size, the difference in prevalence among lineages was larger when imputed lineages were used (**Fig 6B**).

**Figure 6.**
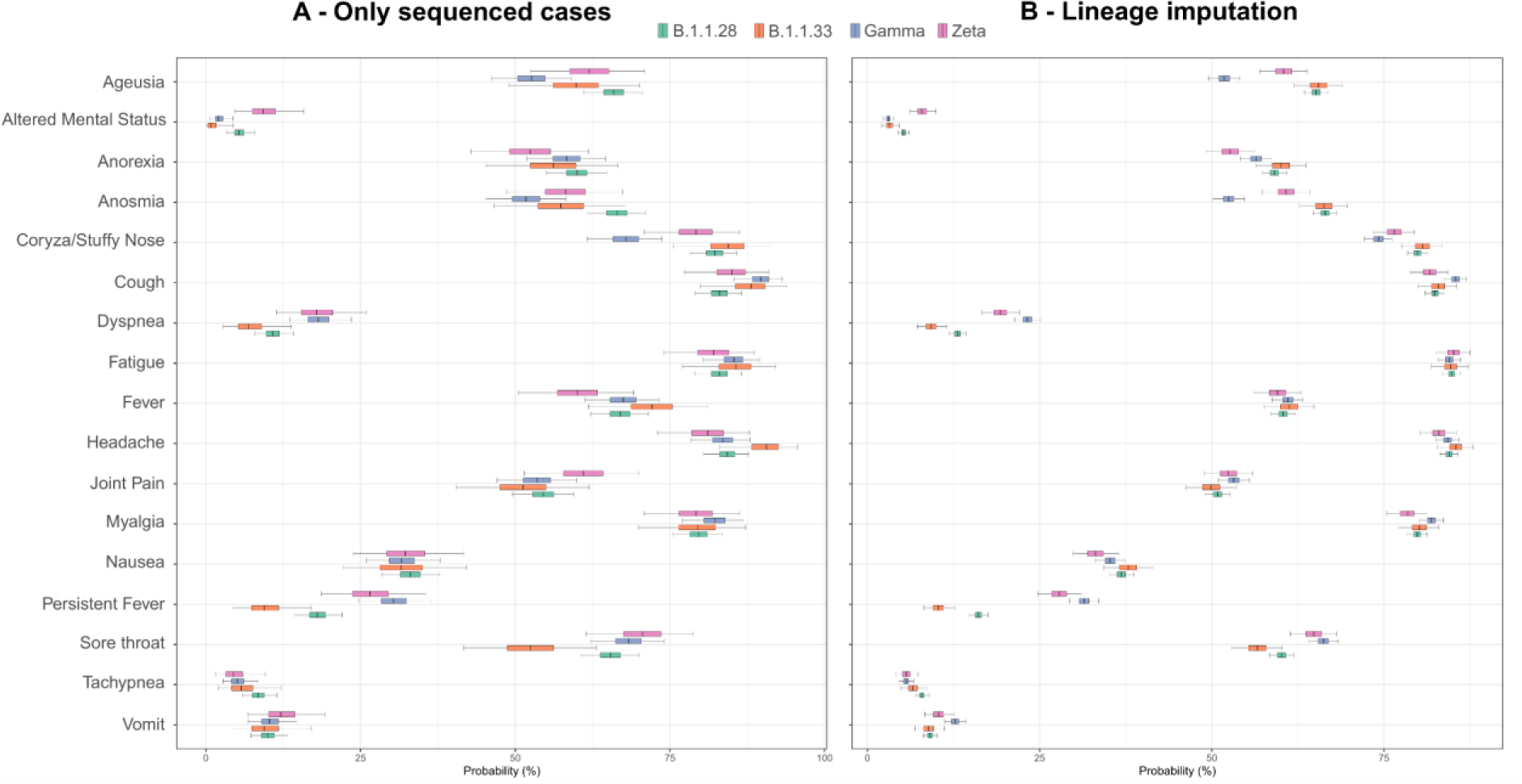
Probability of an individual infected with a given lineage reporting a given symptom in any of the visits. In (A), only the 879 sequenced PCR+ cases were considered in this analysis, while in (B) all PCR+ cases were considered. In (B), non-sequenced PCR+ cases had their corresponding lineage imputed based on the prevalence of each lineage at the date of onset (see Methods for detailed description on the imputation procedure).

To better understand which symptoms are more frequent, we performed one hypothesis test for each symptom and VOC/VOI and used Benjamini-Hochberg correction to adjust for the large number of tests (**Fig. 4S**, **Table 2S**). The Benjamini-Hochberg correction is conservative as it assumes hypothesis tests are independent, while symptoms caused by a given lineage may be correlated. For this reason, we also highlight p-values below the 0.05 threshold in **Fig. 4S**, and we estimated the probability of a given symptom being more frequently caused by Gamma or Zeta when compared to B.1.1.28 and B.1.1.33 (**Fig. 5S**). To further validate our results, we also displayed symptom prevalence across age groups and study phases (**Figure 6S**). Disease severity as measured by dyspnea and persistent fever was higher during the second and third phase (dominated respectively by Zeta and Gamma) for all adult age strata, and anosmia and ageusia were smaller during the third phase. Altered mental status was more common during the second phase for all age groups, but the difference in prevalence across phases was smaller in 0-19 and 80+ age ranges. Note the null hypothesis that the prevalence of altered mental status is the same for Zeta and B.1.1.28 or B.1.1.33 was not rejected (**Table 2S**, **Fig. 4S**), as the p-value for the hypothesis test is 0.08, even though **Fig. 6** and **Fig. 6S** suggest this symptom is more prevalent for Zeta. This is because altered mental status is also more prevalent in B.1.1.28 when compared to B.1.1.33 although less than in Zeta. Thus, grouping B.1.1.28 and B.1.1.33 attenuates the difference in prevalence for Zeta, increasing the p-value. For this reason, we performed six additional hypothesis tests for altered mental status comparing Zeta, B.1.1.28 and B.1.1.33 separately (**Table 3S**), confirming the larger prevalence of altered mental status for Zeta and B.1.1.28.

## 4. Discussion

We analysed the data obtained from a web based platform created to provide care to COVID-19 cases in São Caetano do Sul (5) that represents 63% of all positive cases notified in the city. In addition to epidemiological and clinical data, PCR positive samples could be retrieved for sequencing allowing a detailed description of the evolution of the epidemic in the city and the comparison of disease presentation among the different lineages. Brazil underwent one of the most significant epidemics in the world, with important regional differences. The epidemic in São Caetano do Sul was similar to the metropolitan region of Sao Paulo with a prolonged and plateau-shaped first wave followed by a more prominent second wave driven initially by Zeta lineage that was surpassed by Gamma VOC in Feb 2021. It was estimated that by Dec 2020, 26.6% of the population in the city of Sao Paulo had been infected (18).

**Fig. 4S, 5S, 6S** and **Table 2S** demonstrate that disease presentation differs among lineages, as Gamma and Zeta were significantly more likely to present more severe symptoms such as dyspnea (1.81 and 1.79 fold) and persistent fever (1.85 and 1.61 fold) when compared to ancestral lineages. In the original work describing the Gamma VOC we suggested that it caused 1.2 to 1.9 times more deaths (6). By evaluating the attack rate in Manaus during the first and second wave using a cohort of blood donors, Prete et al. (17) concluded that Infection Hospitalisation Rate (IHR) and Infection Fatality Rate (IFR) were higher during the Gamma-dominated period.

Anosmia and ageusia were less frequent among patients infected with the Gamma VOC, which was also reported by Luna-Muschi et al (2) in a cohort of health care workers, while mental related symptoms were more present among Zeta VOI infected cases. Interestingly, Zeta infection presented more neurological symptoms, which was not previously reported.

Additionally, during the period when Gamma was responsible for the majority of cases, the case fatality rate (CFR), hospitalisation rate (CHR), and in-hospital fatality rate (HFR) were higher. Banho et al. (19) evaluated the rate of severe and non-severe cases in the city of São José dos Campos in Brazil and detected a higher proportion of severe cases especially among the younger population after the introduction of the Gamma variant. Despite the more severe symptoms associated with Zeta infection, there was no significant increase in the hospitalisation rate during the period. Even though increased intrinsic severity of the disease may be associated with larger CFR and CHR, it does not necessarily lead to an increased HFR, as disease severity depends on the risk of hospitalisation. This effect is studied in (3), where a model-based analysis of data from hospitalised patients from Brazil showed that in-hospital fatality rates during the Gamma-dominated wave in Brazil were primarily associated with geographic inequities and shortages in healthcare capacity rather than with the Gamma VOC.

Our study demonstrates that many valuable insights regarding disease transmission and symptom manifestation can only be attained by integrating epidemiological, clinical, and genomic data. The establishment of an online platform to collect data associated with routine care was essential in enabling the acquisition of large, unbiased data and samples.

## Data Availability

Anonymized data and code are used to generate the main analyses of this work are available at https://github.com/carlosprete/coronasaocaetano. This link also contains the ID of all samples submitted to GISAID for this study.

## Author Contributions

**Conception**: ECS, NRF **Collection and processing of samples:** FCSS, ERM, MCMC, FEL, IMC, BAO, HGOP, IMC, CASM, TC **Investigation**: FCSS, CAPJ, IMC, ERM, LC, LFB **Analysis:** FCSS, CAPJ, VHN, KVP, LA, DSC, FRRM, ECS, NRF. **Interpretation:** FCSS, CAPJ, LA, IMC, KVP, SFC, KVP, VHN, ECS, NRF **Drafting:** FCSS, CAPJ, IMC, SFC, VHN, ECS, NRF. **Revising:** All authors.

## Supplementary Material

**Figure 1S.**
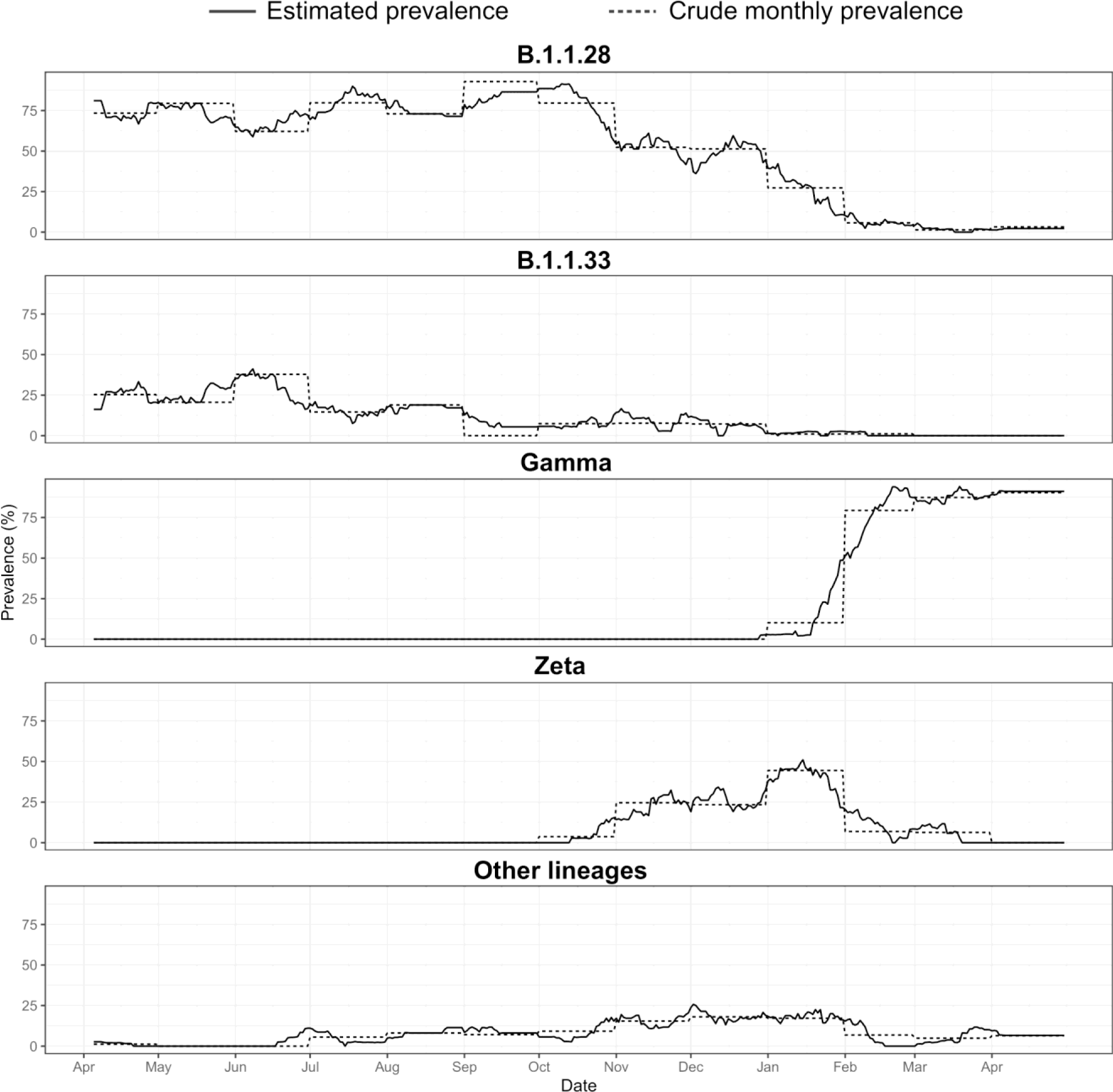
Estimated lineage prevalence compared with the crude monthly lineage prevalence, defined as the ratio between the number of samples of a given sequence in each month and the total number of samples in the same month.

**Figure 2S.**
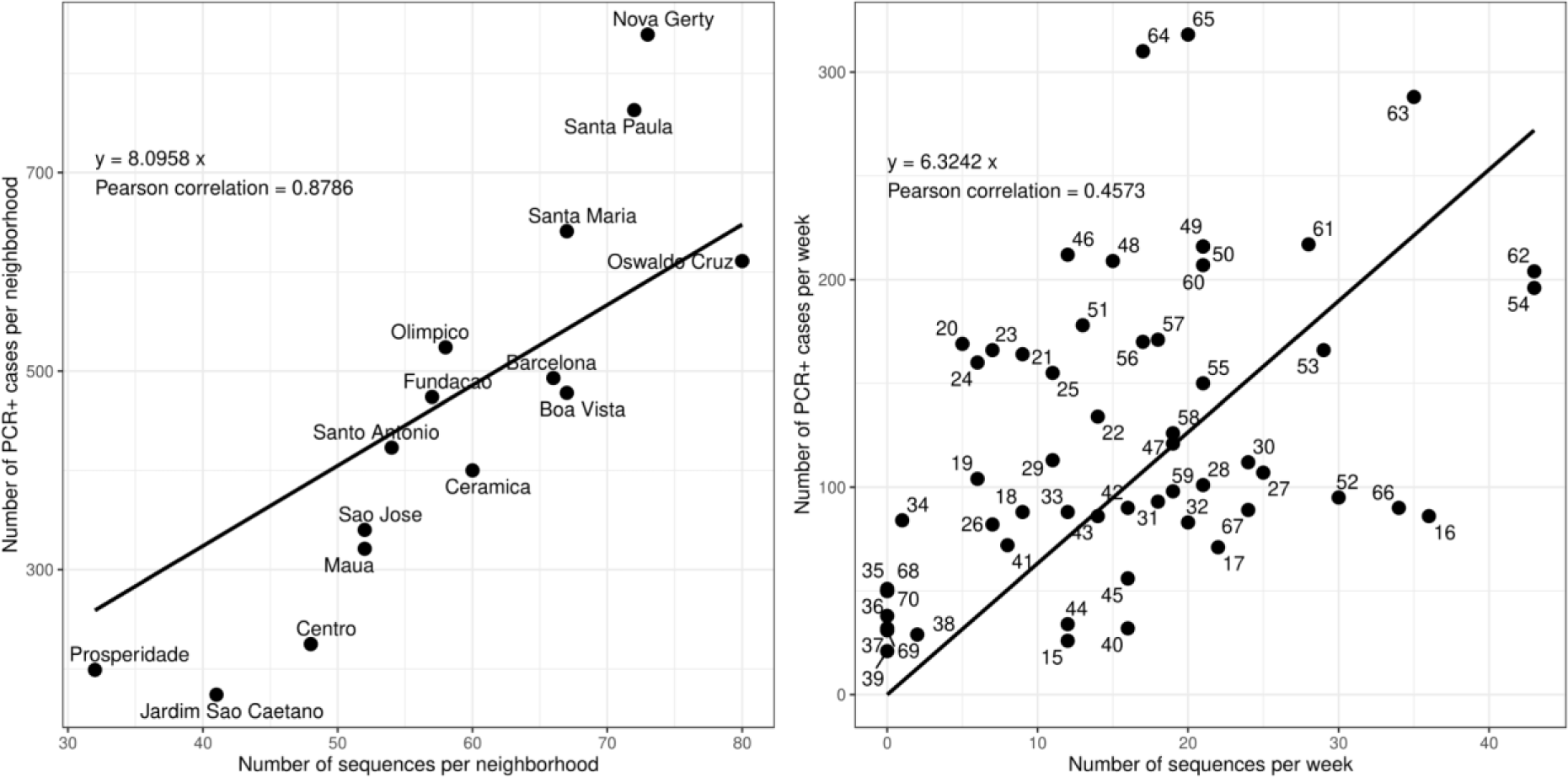
Pearson’s Correlation Chart demonstrating that the number of sequences in our dataset strongly correlates with confirmed cases of SARS CoV-2 PCR spatially (9A) and temporally (B) (Pearson correlation = 0.87 and 0.45, respectively). The line is the linear function (with no intercept) that best fits the data points in the mean square sense.

**Figure 3S.**
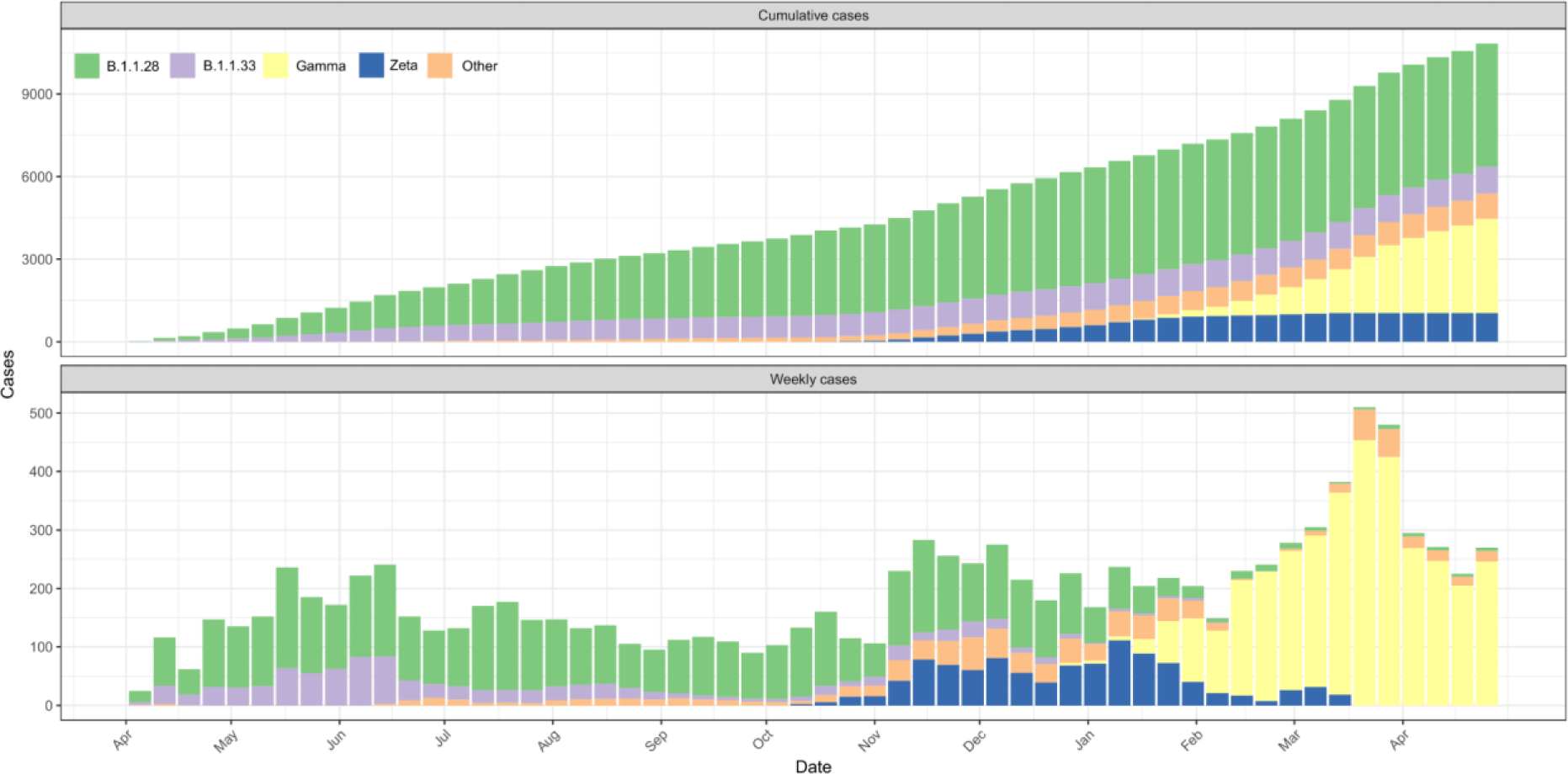
Estimates for cumulative and weekly cases caused by each lineage in São Caetano do Sul. By April 30 2021, 41.3% of the cases were caused by B.1.1.28, followed by Gamma (31.7%), Zeta (9.6%) and B.1.1.33 (9.0%).

**Figure 4S.**
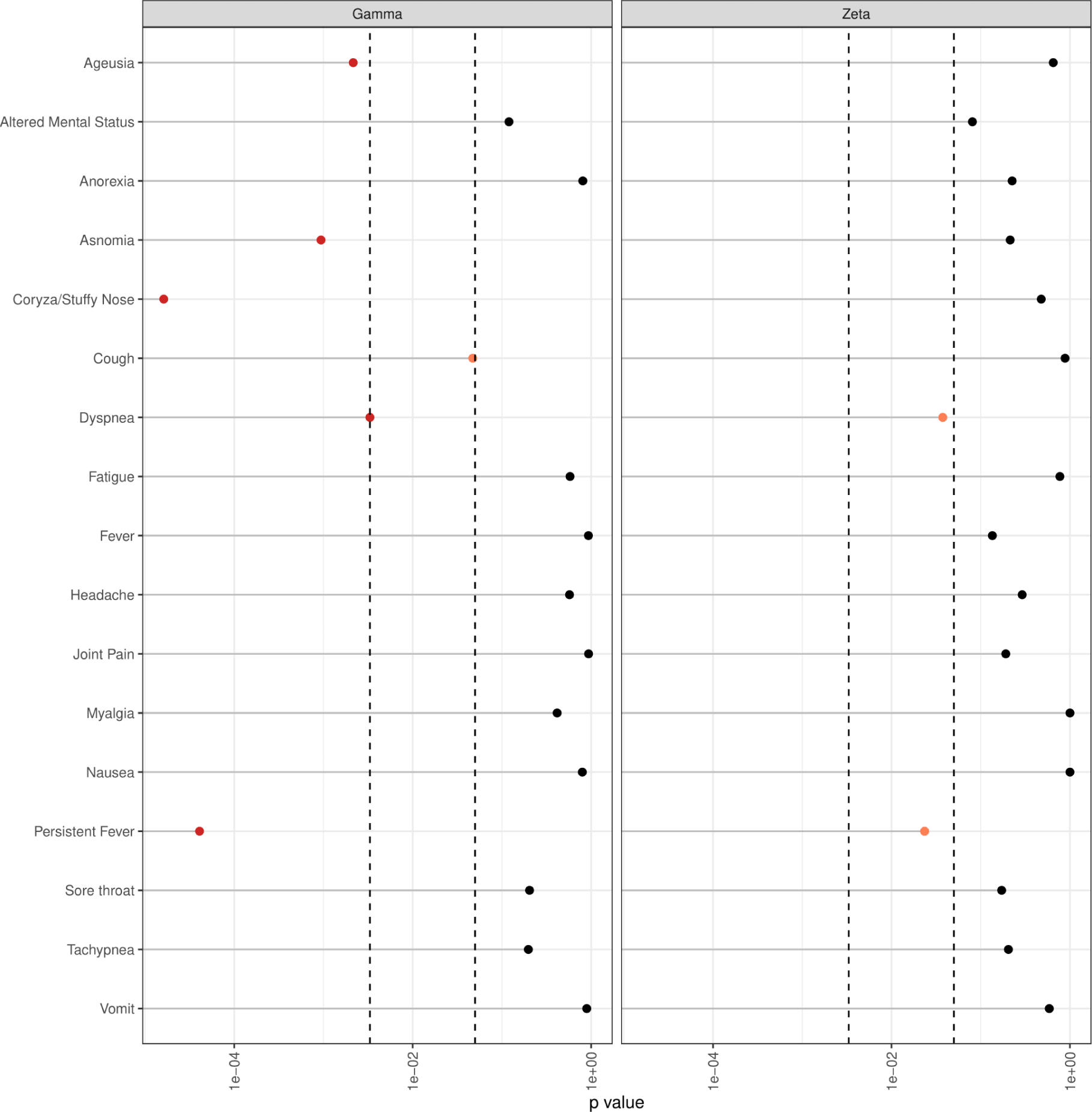
P-values obtained by testing if patients infected by a given VOC or VOI (Gamma or Zeta) is more likely to cause a given symptom than B.1.1.28 and B.1.1.33. We used the exact Fisher test with a confidence level of 95% (points in red or orange). Since 34 hypothesis tests were performed, we performed a Benjamini-Hochberg correction with a false positive rate of 10%, which rejected all hypothesis tests with p <= 0.0033 (in red).

**Figure 5S.**
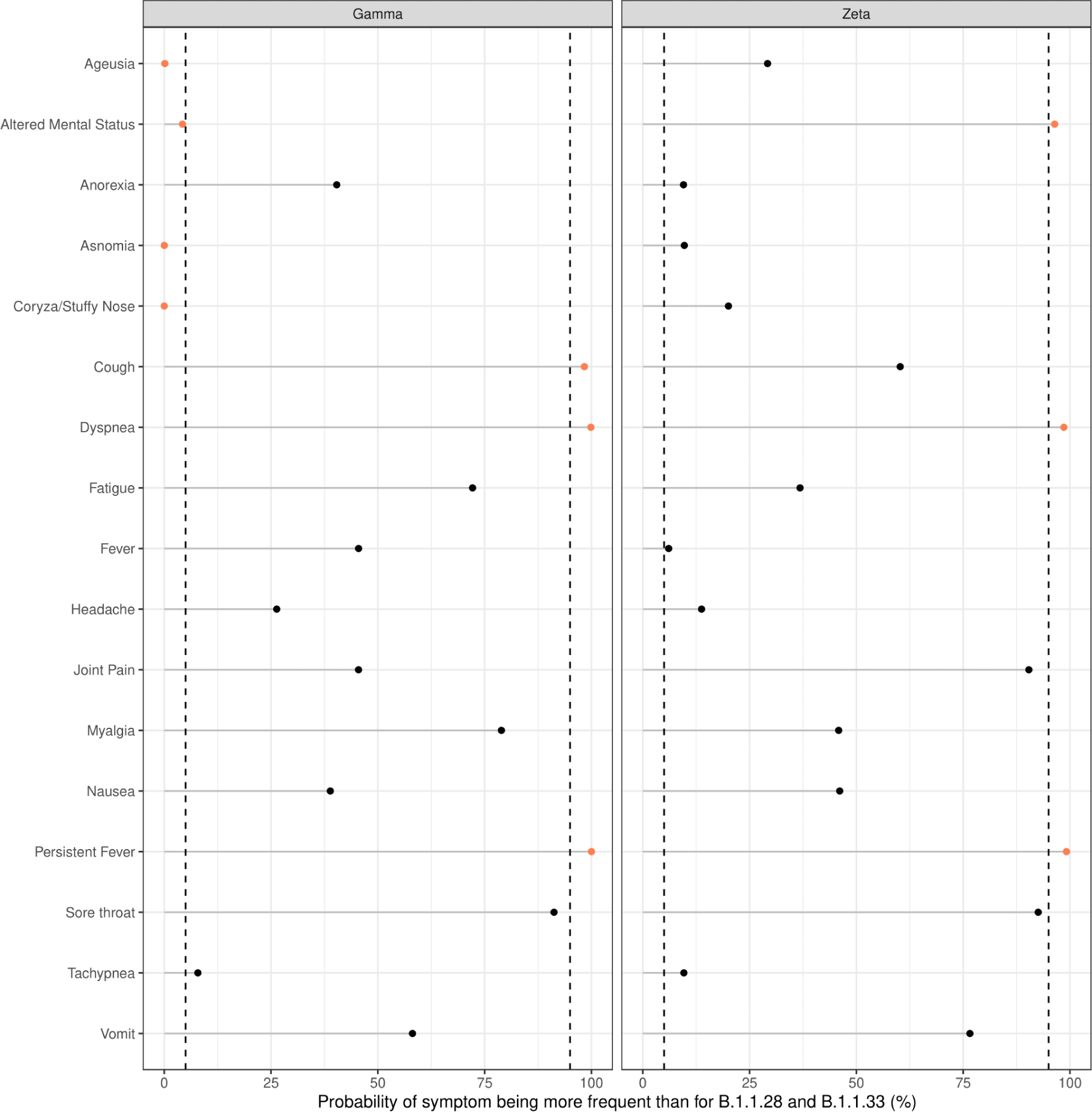
Probability of a given symptom being more frequently caused by Gamma or Zeta when compared to B.1.1.28 and B.1.1.33. Dashed lines represent probabilities of 5% and 95%, and red points represent symptoms with more extremal probabilities than these thresholds. For instance, ageusia, altered mental status, anosmia and coryza/stuffy nose are more likely to occur in patients infected by B.1.1.28 and B.1.1.33 than by Gamma. On the other hand, patients infected by Gamma are more likely to present cough, dyspnea and persistent fever than patients infected by B.1.1.28 and B.1.1.33. See methods for details on how probabilities were calculated

**Figure 6S.**
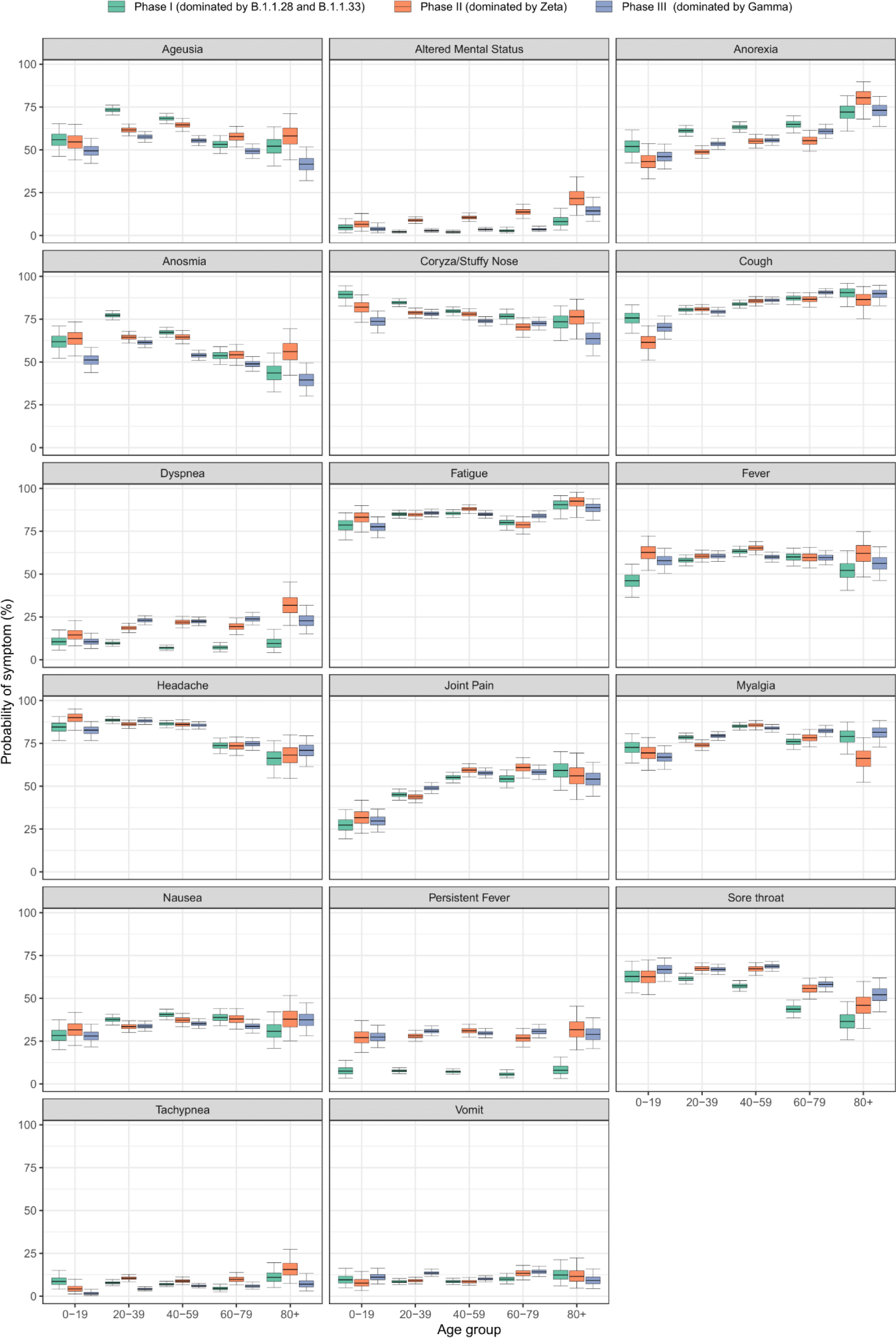
Probability of occurrence of a given symptom in terms of the age group and phase of the study (I, II, III). Boxes show the median and 50% credible intervals, and whiskers indicate the 95% credible intervals. Warning signs for severity: fever/persistent fever, dyspnea/difficulty breathing and tachypnea, altered mental status.

**Table 1S.** Main SARS-CoV-2 lineage groups identifier.

| SARS-CoV-2 Lineages |  |  |  |  |  |  | Phase |
| --- | --- | --- | --- | --- | --- | --- | --- |
| Month/Year | B.1.1.28 | B.1.1.33 | Others* | Gamma ** | Zeta | Total |  |
| April/2020 | 59 | 19 | 1 |  |  | 79 | Phase 1 |
| May/2020 | 59 | 10 | 4 |  | 1 | 74 |  |
| Jun/2020 | 29 | 16 | 1 |  |  | 46 |  |
| July/2020 | 37 | 6 | 2 |  |  | 45 |  |
| Aug/2020 | 22 | 7 | 3 |  |  | 32 |  |
| Sept/2020 | 13 |  | 1 |  |  | 14 |  |
| Oct/2020 | 43 | 4 | 5 |  | 2 | 54 | Phase 2 |
| Nov/2020 | 34 | 5 | 10 |  | 16 | 65 |  |
| Dez/2020 | 57 | 7 | 21 |  | 26 | 111 |  |
| Jan/2021 | 27 | 1 | 17 | 16 | 43 | 104 | Phase 3 |
| Feb/2021 | 5 | 1 | 6 | 69 | 6 | 87 |  |
| Mar/2021 | 2 | 1 | 6 | 124 | 9 | 142 |  |
| April/2021 | 1 |  | 3 | 22 |  | 26 |  |
| **Total** | **388** | **77** | **80** | **231** | **103** | **879** |  |

\* Other: B.1.1, B.1.1.174, B.1.1.348, B.1.1.135, B.1.1.371, B.1.1.274, B.1.1.397, N9, P7, B1.1.7, B.1.1.10.
\*\* Gamma group: P.1, P.1.10, P.1.14.

**Table 2S.** Relative risk of the occurrence of a given symptom using as reference the prevalence of that symptom for lineages B.1.1.28 and B.1.1.33. Benjamini-Hochberg correction with a false positive rate of 10% leads to rejection of all hypothesis tests with p < 0.0033. An asterisk was added after the p-values to indicate the hypothesis test was rejected.

|  | Gamma vs B.1.1.28 and B.1.1.33 |  | Zeta vs B.1.1.28 and B.1.1.33 |  |
| --- | --- | --- | --- | --- |
| Symptom | Relative Risk (95% CrI) | p-value | Relative Risk (95% CrI) | p-value |
| Ageusia | 0.81 (0.70 - 0.93) | 2.15E-03 * | 0.96 (0.80 - 1.11) | 6.49E-01 |
| Altered Mental Status | 0.46 (0.15 - 1.12) | 1.20E-01 | 2.09 (0.95 - 4.17) | 8.07E-02 |
| Anorexia | 0.98 (0.86 - 1.11) | 8.06E-01 | 0.88 (0.71 - 1.06) | 2.25E-01 |
| Anosmia | 0.80 (0.69 - 0.91) | 9.38E-04 * | 0.90 (0.74 - 1.06) | 2.14E-01 |
| Coryza/Stuffy Nose | 0.82 (0.74 - 0.90) | 1.62E-05 * | 0.96 (0.85 - 1.05) | 4.76E-01 |
| Cough | 1.07 (1.00 - 1.13) | 4.71E-02 | 1.01 (0.91 - 1.10) | 8.81E-01 |
| Dyspnea | 1.81 (1.22 - 2.66) | 3.32E-03 | 1.79 (1.07 - 2.83) | 3.75E-02 |
| Fatigue | 1.02 (0.95 - 1.09) | 5.79E-01 | 0.98 (0.88 - 1.07) | 7.71E-01 |
| Fever | 0.99 (0.89 - 1.10) | 9.31E-01 | 0.89 (0.74 - 1.03) | 1.35E-01 |
| Headache | 0.98 (0.91 - 1.04) | 5.72E-01 | 0.95 (0.85 - 1.04) | 2.91E-01 |
| Joint Pain | 0.99 (0.85 - 1.14) | 9.35E-01 | 1.13 (0.94 - 1.33) | 1.91E-01 |
| Myalgia | 1.03 (0.95 - 1.11) | 4.15E-01 | 0.99 (0.88 - 1.10) | 1.00E+00 |
| Nausea | 0.97 (0.77 - 1.22) | 7.96E-01 | 0.99 (0.71 - 1.32) | 1.00E+00 |
| Persistent Fever | 1.85 (1.40 - 2.45) | 4.08E-05 * | 1.61 (1.08 - 2.35) | 2.35E-02 |
| Sore throat | 1.08 (0.96 - 1.21) | 2.04E-01 | 1.12 (0.96 - 1.28) | 1.72E-01 |
| Tachypnea | 0.64 (0.33 - 1.18) | 1.98E-01 | 0.57 (0.20 - 1.30) | 2.04E-01 |
| Vomit | 1.05 (0.64 - 1.66) | 8.92E-01 | 1.23 (0.65 - 2.15) | 5.86E-01 |

**Table 3S.** Relative Risks and p-values for the occurrence of altered mental status. In each column, the second lineage displayed is used as reference for calculating relative risks. A Benjamini-Hochberg correction with a false positive rate of 10% leads to rejection of all hypothesis tests with p < 0.050. An asterisk was added after the p-values to indicate the hypothesis test was rejected.

|  | **Zeta vs**<br>**B.1.1.28** | **Zeta vs**<br>**B.1.1.33** | **Zeta vs**<br>**Gamma** | **B.1.1.28**<br>**vs**<br>**B.1.1.33** | **Gamma**<br>**vs**<br>**B.1.1.28** | **Gamma**<br>**vs**<br>**B.1.1.33** |
| --- | --- | --- | --- | --- | --- | --- |
| **Relative**<br>**Risk (95%**<br>**Crl)** | 1.75 (0.80 - 3.51) | 11.26 (1.79 - 302.6) | 4.55 (1.64 - 14.71) | 6.42 (1.16 - 186.29) | 0.38 (0.12 - 0.94) | 2.33 (0.32 - 73.82) |
| **p-value** | 0.2375 | 0.0053* | 0.0045* | 0.0327* | 0.0500* | 0.5759 |

## Bibliography

1. Gonzalez-Reiche AS, Hernandez MM, Sullivan MJ, Ciferri B, Alshammary H, Obla A, et al. Introductions and early spread of SARS-CoV-2 in the New York City area. Science. 2020 Jul 17;369(6501):297–301.

2. Luna-Muschi A, Borges IC, de Faria E, Barboza AS, Maia FL, Leme MD, et al. Clinical features of COVID-19 by SARS-CoV-2 Gamma variant: A prospective cohort study of vaccinated and unvaccinated healthcare workers. J Infect. 2022 Feb;84(2):248–88.

3. Brizzi A, Whittaker C, Servo LMS, Hawryluk I, Prete CA, de Souza WM, et al. Spatial and temporal fluctuations in COVID-19 fatality rates in Brazilian hospitals. Nat Med. 2022 Jul;28(7):1476–85.

4. Nonaka CKV, Gräf T, Barcia CA de L, Costa VF, de Oliveira JL, Passos R da H, et al. SARS-CoV-2 variant of concern P.1 (Gamma) infection in young and middle-aged patients admitted to the intensive care units of a single hospital in Salvador, Northeast Brazil, February 2021. Int J Infect Dis. 2021 Oct;111:47–54.

5. Leal FE, Mendes-Correa MC, Buss LF, Costa SF, Bizario JCS, de Souza SRP, et al. Clinical features and natural history of the first 2073 suspected COVID-19 cases in the Corona São Caetano primary care programme: a prospective cohort study. BMJ Open. 2021 Jan 12;11(1):e042745.

6. Faria NR, Mellan TA, Whittaker C, Claro IM, Candido D da S, Mishra S, et al. Genomics and epidemiology of the P.1 SARS-CoV-2 lineage in Manaus, Brazil. Science. 2021 May 21;372(6544):815–21.

7. Buss LF, Prete CA, Abrahim CMM, Mendrone A, Salomon T, de Almeida-Neto C, et al. Three-quarters attack rate of SARS-CoV-2 in the Brazilian Amazon during a largely unmitigated epidemic. Science. 2021 Jan 15;371(6526):288–92.

8. Candido DS, Claro IM, de Jesus JG, Souza WM, Moreira FRR, Dellicour S, et al. Evolution and epidemic spread of SARS-CoV-2 in Brazil. Science. 2020 Sep 4;369(6508):1255–60.

9. Instituto Brasileiro de Geografia e Estatístic. IBGE [Internet]. [cited 2023 Jun 21]. Available from: https://www.ibge.gov.br/cidades-e-estados/sp/sao-caetano-do-sul.html

10. Quick J, Grubaugh ND, Pullan ST, Claro IM, Smith AD, Gangavarapu K, et al. Multiplex PCR method for MinION and Illumina sequencing of Zika and other virus genomes directly from clinical samples. Nat Protoc. 2017 Jun;12(6):1261–76.

11. nCoV-2019 sequencing protocol v3 (LoCost) [Internet]. [cited 2022 Jun 29]. Available from: https://www.protocols.io/view/ncov-2019-sequencing-protocol-v3-locost-bp2l6n26rgqe/v3

12. Tyson JR, James P, Stoddart D, Sparks N, Wickenhagen A, Hall G, et al. Improvements to the ARTIC multiplex PCR method for SARS-CoV-2 genome sequencing using nanopore. BioRxiv. 2020 Sep 4;

13. Li H. Minimap2: pairwise alignment for nucleotide sequences. Bioinformatics. 2018 Sep 15;34(18):3094–100.

14. Li H, Handsaker B, Wysoker A, Fennell T, Ruan J, Homer N, et al. The Sequence Alignment/Map format and SAMtools. Bioinformatics. 2009 Aug 15;25(16):2078–9.

15. Buss L, Prete CA, Whittaker C, Salomon T, Oikawa MK, Pereira RHM, et al. Predicting SARS-CoV-2 Variant Spread in a Completely Seropositive Population Using Semi-Quantitative Antibody Measurements in Blood Donors. Vaccines (Basel). 2022 Aug 31;10(9).

16. Parag KV. Improved estimation of time-varying reproduction numbers at low case incidence and between epidemic waves. PLoS Comput Biol. 2021 Sep 7;17(9):e1009347.

17. Prete CA, Buss L, Dighe A, Porto VB, da Silva Candido D, Ghilardi F, et al. Serial interval distribution of SARS-CoV-2 infection in Brazil. J Travel Med. 2021 Feb 23;28(2).

18. Prete CA, Buss LF, Whittaker C, Salomon T, Oikawa MK, Pereira RHM, et al. SARS-CoV-2 antibody dynamics in blood donors and COVID-19 epidemiology in eight Brazilian state capitals: A serial cross-sectional study. eLife. 2022 Sep 22;11.

19. Banho CA, Sacchetto L, Campos GRF, Bittar C, Possebon FS, Ullmann LS, et al. Impact of SARS-CoV-2 Gamma lineage introduction and COVID-19 vaccination on the epidemiological landscape of a Brazilian city. Commun Med (London). 2022 Apr 13;2:41.

